# How disease risk awareness modulates transmission: coupling infectious disease models with behavioral dynamics

**DOI:** 10.1101/2021.04.13.21255395

**Authors:** Jaime Cascante-Vega, Samuel Torres-Florez, Juan Cordovez, Mauricio Santos-Vega

## Abstract

Epidemiological models often assume that individuals do not change their behavior or that those aspects are implicitly incorporated in parameters in the models. Typically these assumption is included in the contact rate between infectious and susceptible individuals. For example models incorporate time variable contact rates to account for the effect of behavior or other interventions than in general terms reduce transmission. However, adaptive behaviors are expected to emerge and to play an important role in the transmission dynamics across populations. Here, we propose a theoretical framework to couple transmission dynamics with behavioral dynamics due to infection awareness. We first model the dynamics of social behavior by using a game theory framework. Then we coupled the model with an epidemiological model that captures the disease dynamics by assuming that individuals are more aware of that epidemiological state (i.e. fraction of infected individuals) and reduces their contacts. Our results from a mechanistic modeling framework show that as individuals increase their awareness the steady-state value of the final fraction of infected individuals in a susceptible-infected-susceptible (SIS) model decreases. We also extend our results to a spatial framework, incorporating a spatially-defined theoretical contact network (social network) and we made the awareness parameter dependent on a global or local contact structure. Our results show that even when individuals increase their awareness of the disease, the spatial structure itself defines the steady state solution of the system, in which more connected networks (networks with random or constant degree distributions) results in a population with no change in their behavior. Our work then shows that explicitly incorporating dynamics about the behavioral response dynamics might significantly change the predicted course of the epidemic and therefore highlights the importance of accounting for this source of variation in the epidemiological models.

**Author summary:** We present a theoretical framework for coupling traditional epidemiological models with a behavioral dynamical model in the form of a game-theoretical setting. Here, individual payoffs are assumed to be coupled with the force of infection (FOI) and the transmission probability, which is proportional to the individuals behavior. Our approach studies the temporal dynamics of a mechanistic epidemiological model (SIS) coupled with a prisoners dilemma framework, then we extended the results to an SIS model implemented on a network (social network) using three types of networks: Scale-free, Watts-Strogatz or small world and grid networks. Our results show that behavior can change the final fraction of infected individuals and the fraction of cooperators or individuals who voluntarily take actions to reduce their transmission in the system. In addition, when the dynamics were studied on a contact network we found that the topology of this network plays an essential role in controlling individuals behavior. Specifically, our results show that as the network gets more connected (i.e. degree distribution is random or uniform (Watts-Strogatz or grid networks respectively) disease spread is faster and therefore individuals are not obligated to cooperate. However, when the dynamics are studied in a scale free contact network, as degree distribution follows a power-law, we show that similarly as the mechanistic ODEs model individuals cooperate so their transmission probability is reduced.

## Introduction

Traditional infectious disease transmission models allocate the population into compartments that captures different disease states, and aim to parametrize the rates of transition between those states in a manner that reflects the underlying biology of the disease [1, 2]. Numerous factors influence transmission and are important to consider or model directly in epidemiological models. On of those is the understanding of the effect of individuals behavior on the disease population dynamics, which has recently been highlighted as a response to reduce contact rates and therefore pathogen transmission across populations [3]. Typically, as a disease spreads in a population, individual behavior can change and therefore infection probability can be reduced or amplified [4–7]. Recently, changes in individual contact rates driven by changes in the has been discussed and explicitly modeled [8, 9]. Although, these models do not incorporate an explicit mechanism by which individuals could modify their behavior. The have demonstrated how behavioral aspects play an essential role in disease dynamics. Over time, behavior is expected to vary as population-level disease awareness is modulated by increases in risk and the proportion of the population that has been infected (e.g., risk awareness might be highest near the peak of the epidemic) [9]. However the impact of behavior-time dynamics in controlling transmission is likely intertwined with other variables that directly impact transmission, and can be estimated in a time-variable contact rate. Recently, COVID-19 have highlighted the importance of sustained social distancing to reduce infection risk within the population so health systems capacity is not saturated [3–5]. As epidemics unfold, individuals amass information provided by public health institutions, concerning the status of an epidemic or are strongly influenced by beliefs of the disease in their population. This heterogeneity in the levels of information gathered in certain communities, could modulate the level of adherence to certain interventions. A classic example is the Ebola outbreak in Sierra Leone in where risk communication played an essential role in controlling transmission due to high infection probability given contact with and infectious individual of this disease [CITE https://www.ncbi.nlm.nih.gov/pmc/articles/PMC5782897/pdf/17-1028.pdf,].

Mechanistic models that aim to explain dynamics of human behavior often rely on a game theory framework, where behavior is represented by the strategy that an individual adopts in a decision-making process [10–13]. However, most coupled epidemiological-game theory models assume static games, and therefore do not capture the dynamics of how individuals change their behavior over time, as an epidemic unfolds [14, 15]. In contrast, evolutionary game theory has been developed precisely to model decision-making as a time-dependent process. By using simple assumptions about how different behavioral strategies generate a payoff (i.e., advantage in evolutionary fitness), they model decisions across individuals and therefore how the adoption of a given strategy changes their frequency in time [16–19]. To date, models that accounts for behavioral dynamics have not been coupled with epidemiological models to address the evolution of behavioral strategies in connections with disease transmission.

Both epidemiological and behavioral dynamics have been studied under space-time couplings resulting in complex dynamics depending on the contact or social network where interactions between individuals take place [7, 20]. This highlights the importance of understanding both coupled and dynamical models in defined social networks where contacts represent interaction between individuals-nodes. Understand how networks topology could modulate disease awareness and ultimately how epidemics unfold [21, 22].

In this work, we introduce a novel framework for coupling both epidemiological and behavioral models. Our framework consists of a traditional epidemiological model [2] and a replicator dynamics model [10, 11].By combining these two approaches we were able to dynamically couple individuals behaviors with transmision intensity. Our work shed lights on the interaction between social and behavioral dynamics affecting the epidemiology of the disease. Specifically, our results show that even when individuals increase the disease awareness, the contact network itself defines the steady state solution of the system. In addition, more connected networks (networks with random or constant degree distributions) results in a population with no change in their behavior. Our work shows that explicitly incorporating responsive behavioral dynamics can significantly change the predicted course of an epidemic and highlights the importance of accounting for this source of variation.

## Materials and methods

We study these coupled epidemiological and social dynamics using two dyfferent models: **(1)** We couple two Ordinary Differential Equations systems (ODEs), one to describe the state transitions in the epidemiological model and the other to describe the replicator dynamics. The epidemiological model is a Susceptible-Infected-Susceptible (SIS) compartmental model, where susceptible individuals in the population are those who are immunologically able to acquire a novel pathogen; infected are those who have acquired the pathogen and are infectious. We assume that the ofrce of infection (FOI) is inversely proportional to the fraction of individuals in the population who cooperate *c* (Here cooperation is understood as the strategy that reduce epidemiological risk but not necessarily provides the highest payoff). Then, following the rationale that individuals who do not cooperate might have a higher FOI, we discount the defector (non-cooperator) payoff by the fraction of infected individuals *I/N* times a parameter that we call the awareness *σ*. **(2)** Our other approach was couple the epidemiological dynamics in an explicit contact network, 𝒢 where nodes represent the individuals and the edges represent the contacts [6]. We then sequentially update the replicator equation within the network. Similar to the coupled ODE models, we assume the FOI is inversely proportional to the number of cooperating individuals however as the network model considers contacts of an individual we attempt to model individual access to the state of infectious individuals in two settings that we call global and local information. This follows the rationale that individuals can be informed of the disease at two multiple spatial scales; they may be aware of the proportion of infected individuals within the entire population (global information), or they may know the state of infection for their neighbors defined as the individuals that the individual have contact with (local information). We study the dynamics in three types of theoretical constructed networks: scale-free, random and grid networks.

### Mechanistic ODEs Model

We consider an SIS (susceptible – infected – susceptible) model (Eq. 4) which is a variation of the Kermack & McKendrick model for diseases without or with short term immunity, which forms the basis of almost all the communicable disease models studied. In our SIS model, the population is divided into two classes, where Susceptible (S) can be infected by those already Infected (I) and subsequently become susceptible again due to lack of immunity [2]. We then model strategic interaction between individuals using the replicator dynamics (RD) a concept from evolutionary game theory where population’s behavioral traits are described using biologically inspired operations such as natural selection [10, 11, 23]. Under this dynamics, the percentage growth rate of cooperators *ċ/c* is equal to the excess of payoff respect to the populations payoff. We set *f*_*c*_ and *f*_*n*_ as the fitness of cooperators and non-cooperators respectively. Therefore, the average fitness can be obtained as 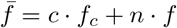

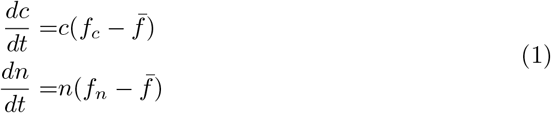

We then model the payoff using the Prisoners Dilemma (PD), an archetypical model displaying the conflict between selfishness and public good [24]. It is a 2 × 2 game with two strategies cooperate or non-cooperate. The payoff modeling pair meetings is defined in Eq. 2. Here we set *S* = 0.5 and *T* = −0.5 as in the Prisoners Dilemma [24]. Note that the row referring to the payoff that receives non-cooperator individuals is discounted by the awareness of the disease *g*(*σ, I*).

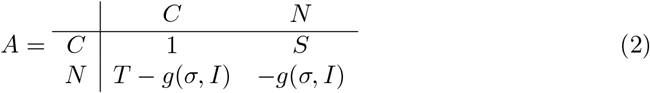

Then the fitness of each strategy *f*_*c*_ and *f*_*n*_ can be written as shown in Eq. 3. Note that the fitness is modeled as the expected payoff given current frequency of cooperators *c* and non-cooperators *n*. Similarly the average fitness 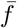 is then the expected given the fitness of each one of the populations.

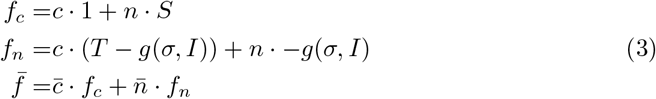

In the typical PD the non-cooperator strategy is the dominating strategy, but the pay-off, if both players defect, is less than the payoff if both cooperate resulting in a social dilemma. Here we assume cooperators payoffs remain the same no matter the FOI in the population, as their behavior might not be perceived less if they follow rules to reduce disease spread. Contrary non-cooperator payoffs reduced proportional to the population consciousness *σ* and the fraction of infected individuals in the population *I/N*. This accounts for the assumption that payoff is reduced proportional to the FOI as is coupled with the number of infected individuals in the population. We then assume individuals strategy affect transmission rate *β* as a function of the fraction of cooperators in the population *c*, then *β*(*c*) = *f* (*c*). As our model attempts to study dynamics in a short-time period (scales at which individuals behavior might change) we do not attempt to include birth or death process in the model. Eq. 4 shows the coupled dynamics resulting in the epi-social model. In SI is included the algebraic reduction of the two dimensional replicator dynamics describing the change of cooperators and non-cooperators to the equation describing the change of cooperators in line *dc/dt*.

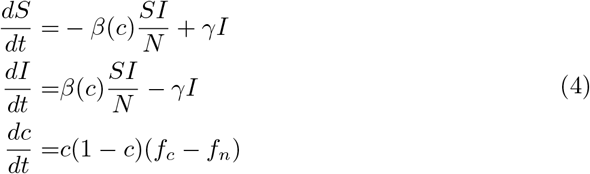

The matrix payoff governing the dynamics of the game are then given by Eq. 2. We assume non-cooperators’ payoff is discounted by the awareness of the disease given the current epidemic state *g*(*σ, I*). The level of consciousness that a population exhibits named *σ*, and the awareness population given the current epidemic state is defined by Eq. 5. Finally, the transmission probability is assumed to be inversely proportional to the fraction of cooperators in the population, one might think in a family of functions for modeling this but by simplicity we only assume contact rate decays exponentially with the cooperator fraction *c* as is shown in the Eq. 6. Additionally this functional response only needs to be parametrized with the nominal contact rate named *β*_max_ in a population where behavior is assumed to not impact transmission at all (i.e. *σ* = 0 an unconscious population).

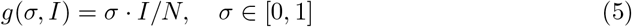

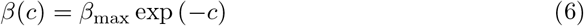

### Network Model

Interactions between individuals (i.e. explicit contacts) are modeled assuming mass-action law; we therefore assume homogeneous contacts between individuals regardless of their state of the disease, susceptible or infected. However, considering explicit social networks give additional insight about the role of contacts, usually studied within theoretical constructed networks [8, 25]. We implemented a SIS model on a network assuming now susceptible individuals get infected by their infected neighbors (individuals with common edges) by a infection probability *β*_*i*_ and recover from the disease with probability *P*_*r*_ = 1*/γ* where *γ* is the recovery rate as used in the ODEs model Eq. 4. Then each individual *i* has a state in the disease named Susceptible *S* or Infected *I* as described by Eq. 7.

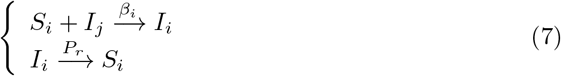

To extend the behavioral dynamics on a network we again use the imitation dynamics update rule given a pay-off matrix as the one described by Eq. 2. We assume that individuals tend to adopt the strategy of more successful neighbors, where success is measured with the payoff as described before, more successful individuals correspond to the ones with greatest payoff in a time step of the simulation. Then successful individuals are pushed to both balance the social dilemma between cooperate or non given the current state of the epidemic. Similar to the replicator dynamics, mean field approximation assumes a two strategy game whose update rule is described by Eq. 8 where the probability of changing from the strategy used by the player *j* to the strategy used by the player *i*, is given by the differences of payoffs of each player *u*_*j*_ and *u*_*i*_ respectively and player irrationality *K*, which we set to 0.5.

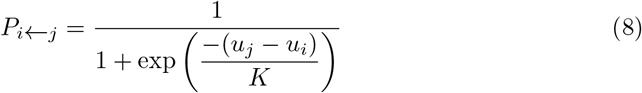

Therefore the payoff of player *i* comes from playing the same symmetric two-player game defined by the used 2 × 2 payoff matrix. In this case her total income can be expressed as Eq. 9.

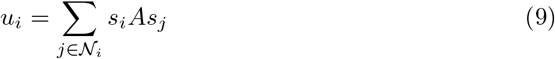

Where *s*_*i*_ is the unit vector describing the the pure strategy played by the individual *i* and the summation runs on the individual neighbors *j* _*i*_ defined by the network structure [17, 20]. The infection rule is given by the infection probability of each node *β*_*i*_, which similarly as the ODEs model this node infection probability will be inversely proportional to the number of cooperating individuals in her neighborhood 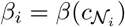. Again for simplicity we only assume that infection probability decays exponentially with the fraction of cooperators in her neigh-boors as is shown in the Eq. 10. This functional response again can only be parametrized using the nominal infection probability named *β*_max_.

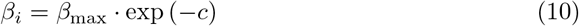

The social dynamics will consequently depend on the current state of the disease. We discount the payoff of non-cooperator individuals by the awareness parameter *σ* and the current state of the disease *g*(*σ, I*). However for modeling the effect on local vs global spread of the information we assume this discount factor will depend on the state of the disease in neighboring nodes (local information) or in the whole population (global information) as described by Eq. 11. This approximation follows the rationality that disease dynamics are governed by local outbreaks (infection among nodes and their neighbors) as is shown in Fig. 1B, C.

**Fig 1.**
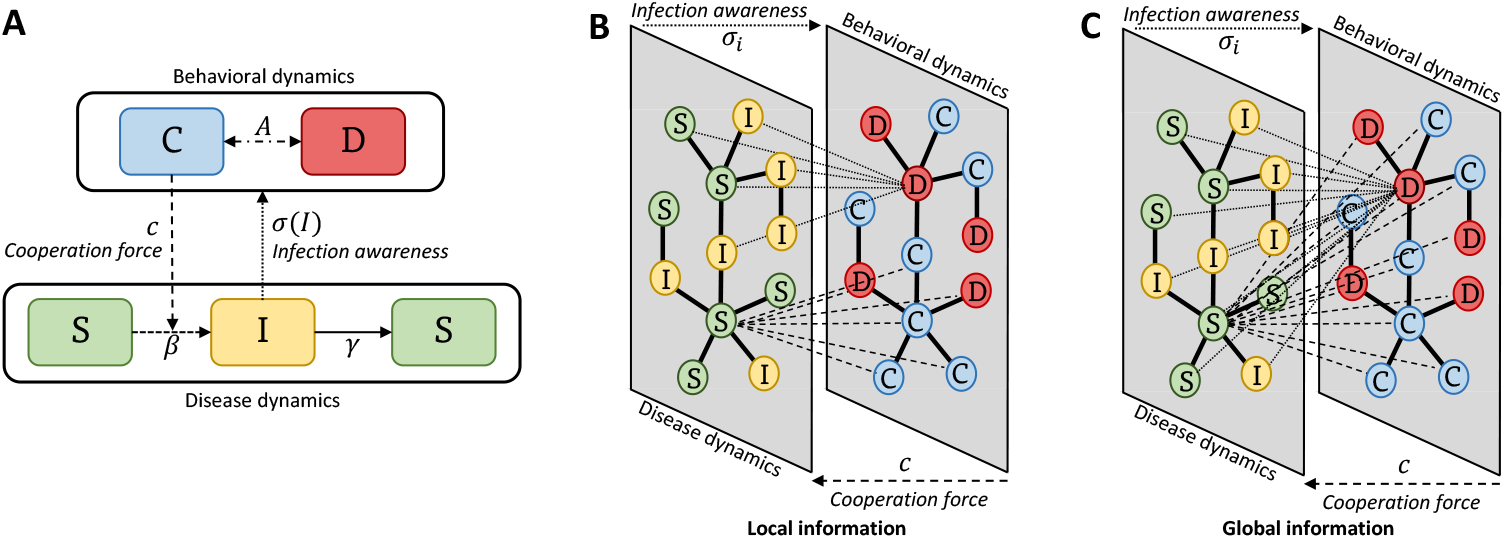
Coupling disease and behavioral dynamics. **A** Schematics for a deterministic framework which is replicated in **B** and **C** aiming for an individual-based approach. **B-C** Schematics for the probabilistic implementation on a multi-layer network. **B** Local information is only transmitted through adjacent nodes Ω_*i*_. For the behavioral dynamics, an individual’s behavioral strategies are only affected by their awareness *σ* of the disease state of the neighboring nodes (omega) at time *t*. Similarly, the infection probability *β*_*i*_ of any node, depends only on the behavior of their neighbors, which is related to the number of cooperating individuals *c*. In contrast, **C** describes global information transmission in the population. Here, individuals base their strategies based on the awareness *s*_*i*_ of the infection state for the entire network. Similarly, the infection probability *β*_*i*_ of any node will depend on the cooperation *c* of all the population.

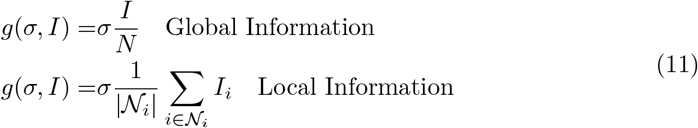

### Simulation and steady state analysis

For both models we assume individuals in average are infectious for 7 days and therefore the recovery rate is *γ* = 1*/*7. We first study the steady state of both ODEs and network models and characterize the dynamics of the system in the steady state. We calculated the fraction of infected individuals *Ī*= *I/N* and the fraction of cooperators *c*. Given that the network model is stochastic and sensitive to the number of infected nodes in the network at the beginning, we ran 100 simulations in a 2000-node network for *T* = 150 days after the first infection was seeded. Additionally, to compute the steady state 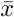 of variable *x*(*t*) we averaged in the last week of the simulation as shown in equation 12 to reduce the effect of the stochastic behavior in the network simulation.

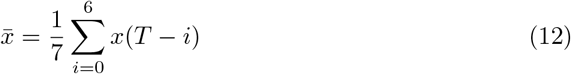

To seed the initial conditions we set the initial condition as 50% of the population as cooperators. Note that as the non-cooperator strategy is the dominant strategy in the payoff matrix this model is not sensible to the initial conditions. This seeding follows the rationale that a randomly-determined fraction of the population cooperates voluntarily toward reducing their risk of getting infected with the disease. We then vary the contact rate or probability of infection *β* and *β*_*i*_ respectively for ODEs and network model between 0.3 and 1. In the results rather than presenting the contact rate *β* we use the basic reproductive number *R*_0_ = *β/γ* of the disease as is a quantity one can relate easily with an specific disease and itself has a meaning in the epidemiological context. This basic reproductive number *R*_0_ correspond to the expected number of infected individuals given an initial one in a completely susceptible or free of the disease population. This results in varying the *R*_0_ between 0.7 and 7 with the specified recovery rate *γ* = 1*/*7.

Finally, to analyze the role of the heterogeneity in contact network favoring cooperation, we study the temporal dynamics of the model at two scales using three levels of awareness that we call full, partial and half awareness for *σ* ∈ {1, 0.7, 0.5} respectively. These scales correspond to a population scale and a clustered-neighbors scale, this idea follows the rationale that networks exhibit a high clustering coefficient [20] suggesting that in steady state hubs of nodes might favor cooperation. We therefore analyze how was the disease dynamics in those clusters. To determine the effect of transmissibility of the disease we consider five values of *R*_0_ ∈ {6.3, 4.2, 3.1, 2.1, 1.5} We do not consider *R*_0_ values less than 1. In SI we include further information of the clustering algorithm used.

## Results

Our results focuses on understanding the role of behavior modeled as the replicator dynamics present in the ODEs model and the imitation update rule in the network model. Figure 2 present the steady state solution for all the models used, Fig 2A shows steady states for the ODEs system using the SIS and replicator dynamics-coupled model, figures on the left correspond to the final fraction of infected individuals running the model with different values of *R*_0_ and consciousness *σ*. Darker red correspond to a higher fraction of infected individuals in the steady state. Similarly, figures on the right show the final fraction of cooperators in the model where green corresponds to a higher final fraction of cooperators in the steady state. Real contact networks are usually highly heterogeneous, and often scale-free as well as highly clustered. Figs 2B, 2C and 2D shows the median steady state dynamics after running the model 100 iterations for scale-free, Watts-Strogatz or small-world and grid graphs respectively. Figs 2B.1, 2C.1 and 2D.1 shows steady state fraction of infected individuals *Ī* = *I/N*. The heat-maps on the left correspond to model runs in the global scenario, where awareness discount factor considers all the infected individuals in the population, while heat-maps on the right correspond to runs of the model in a local scenario, where awareness discount factor depends on each individual proportional to the infected fraction in their neighborhood. Similarly, Figs 2B.2, 2C.2 and 2D.2 shows the steady state fraction of cooperating individuals 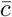. Left and right columns correspond to running the model in the global and local setting, again these settings affect the awareness discount factor given the current state of the disease.

**Fig 2.**
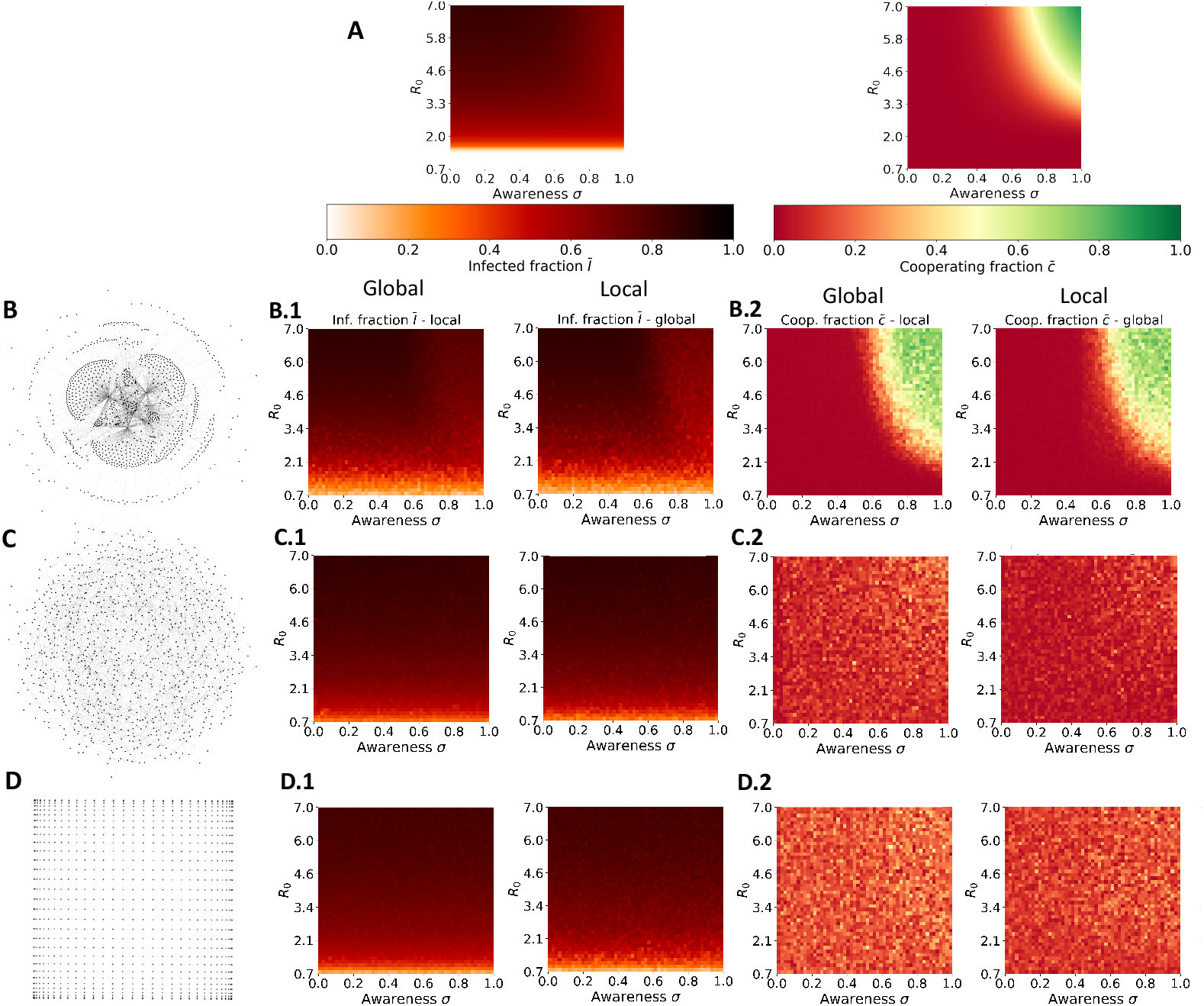
Steady state solution of population infected fraction, and cooperators fraction. Local and Global simulations are only presented for Network Models. **A)** ODEs model. Network model on Scale-Free graph. **C)** Network model on Watts Strogatz/small-world graph. **D)** Network model on grid graph. The y-axis of all heat-maps represent basic reproductive number *R*_0_ and the x-axis represent awareness level *σ*.

While steady states characterize the overall effect of transmission and social parameters *R*_0_ and *σ* respectively. Temporal dynamics of both infected and cooperators exhibit interesting changing behavior as transmissibility increased, leading the dynamical system to their steady state faster [2, 26]. Then suggesting behavior dynamics, i.e. cooperation will also be faster as a response of reducing the transmission of the disease. We then study time dynamics under three levels of consciousness named full, partial and medium consciousness as we indicated in the methods section and three levels of transmission that relates pathogens from normal transmission ∼2 or highly transmitted ∼6. Specifically we use *R*_0_ ∈ {2.1, 4.2, 6.3} this is shown in Fig. 3. Each row represents a different transmission *R*_0_ value and colors represent a given level of consciousness.

**Fig 3.**
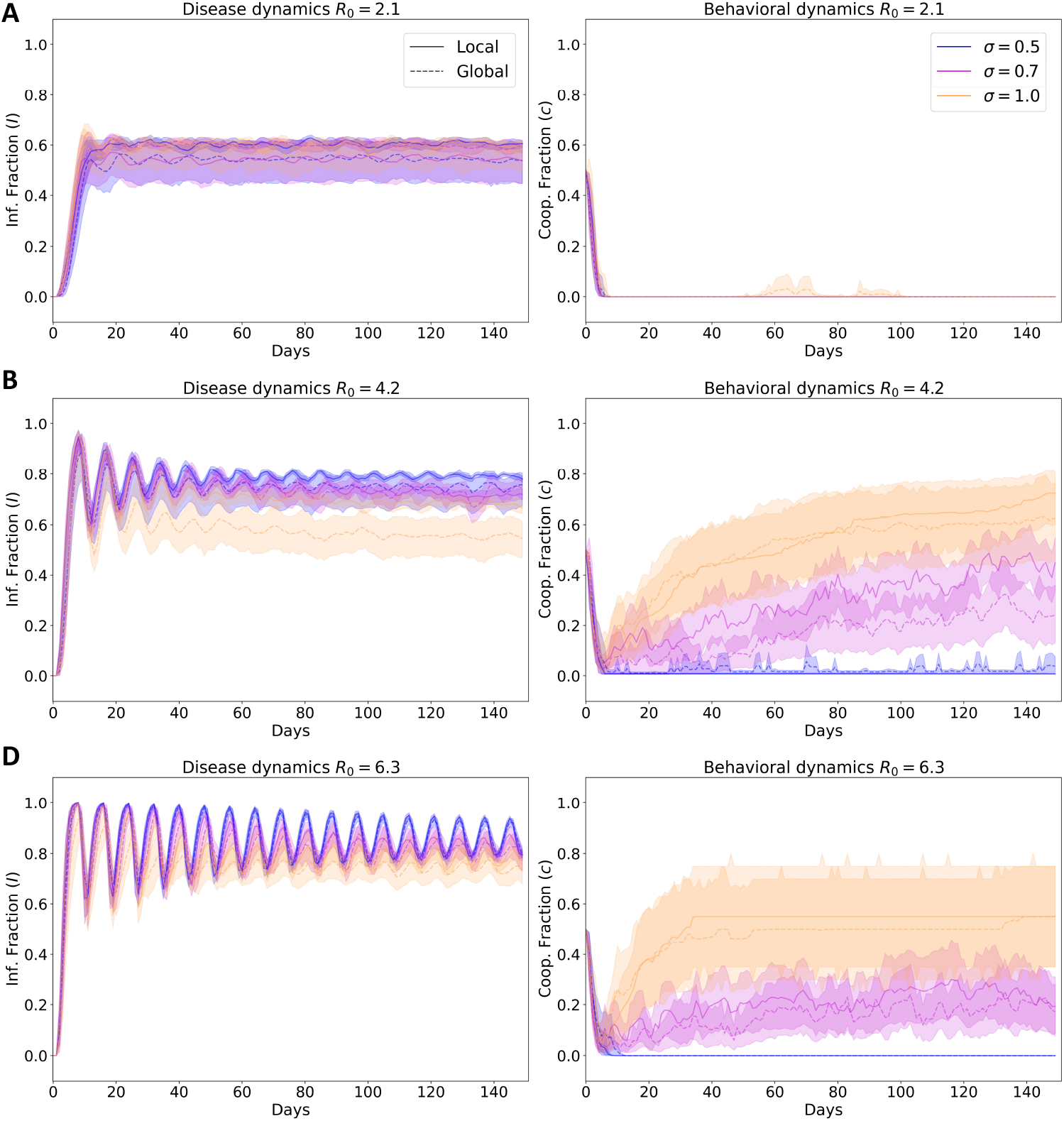
Time dynamics for different levels of consciousness *σ* ∈ {0.5, 0.7, 1.0} and three different levels of transmission **A)** *R*_0_ = 6.3, **B)** *R*_0_ = 4.2, **C)** *R*_0_ = 2.1. For all lines represent the median of the 100 simulations and ribbons represent the 90% quantiles. Left column shows the infected fraction in time *I*(*t*)*/N* and the right column shows the cooperators fraction *c*. The solid line corresponds to running the network model in the global setting while dashed line corresponds to the local setting. We simulate the model for *T* = 150 days after the first infection was seeded.

It has been shown that evolutionary dynamics on scale-free networks might favor cooperation (sustain it in time) in highly connected nodes [20]. Therefore, we hypothesize that if both infected fraction and cooperators fraction might change across scale-free network clusters. In consequence we search for clusters using a node clustering algorithm on the network and plot the fraction of infected individuals; (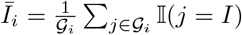 where *𝒢* _*i*_ correspond to the set of nodes identified in the *i* - *th* cluster and 𝕀 (*j* = *I*) = 1 if the *j* - *th* node is infected). Similarly, we computed the cooperator fractions in these clusters 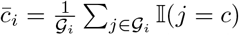, where *𝒢* _*i*_ corresponds to the set of nodes identified in the *i* – *th* cluster and 𝕀(*j* = *c*) = 1 if the *j* - *th* node is cooperating. Fig. 4 shows the temporal dynamics for *T* = 150 days after the seeded index case. The clusters computed are also shown at top left side of Fig 4; note that we only consider the three biggest hubs (top three clusters detected with more nodes). We show the temporal dynamics in Fig. 4B, C, here the row **B** correspond to a maximum level of awareness and the row **C** to a medium level of awareness, we do not show results for low level of awareness as is shown in Fig. 2B.2 cooperation does not occur at that level. The middle and right columns show final infected and cooperators fraction in each cluster; the lines indicate the setting in which we run the simulation, with the solid line corresponding to the global setting and the dashed line corresponding to the local setting.

**Fig 4.**
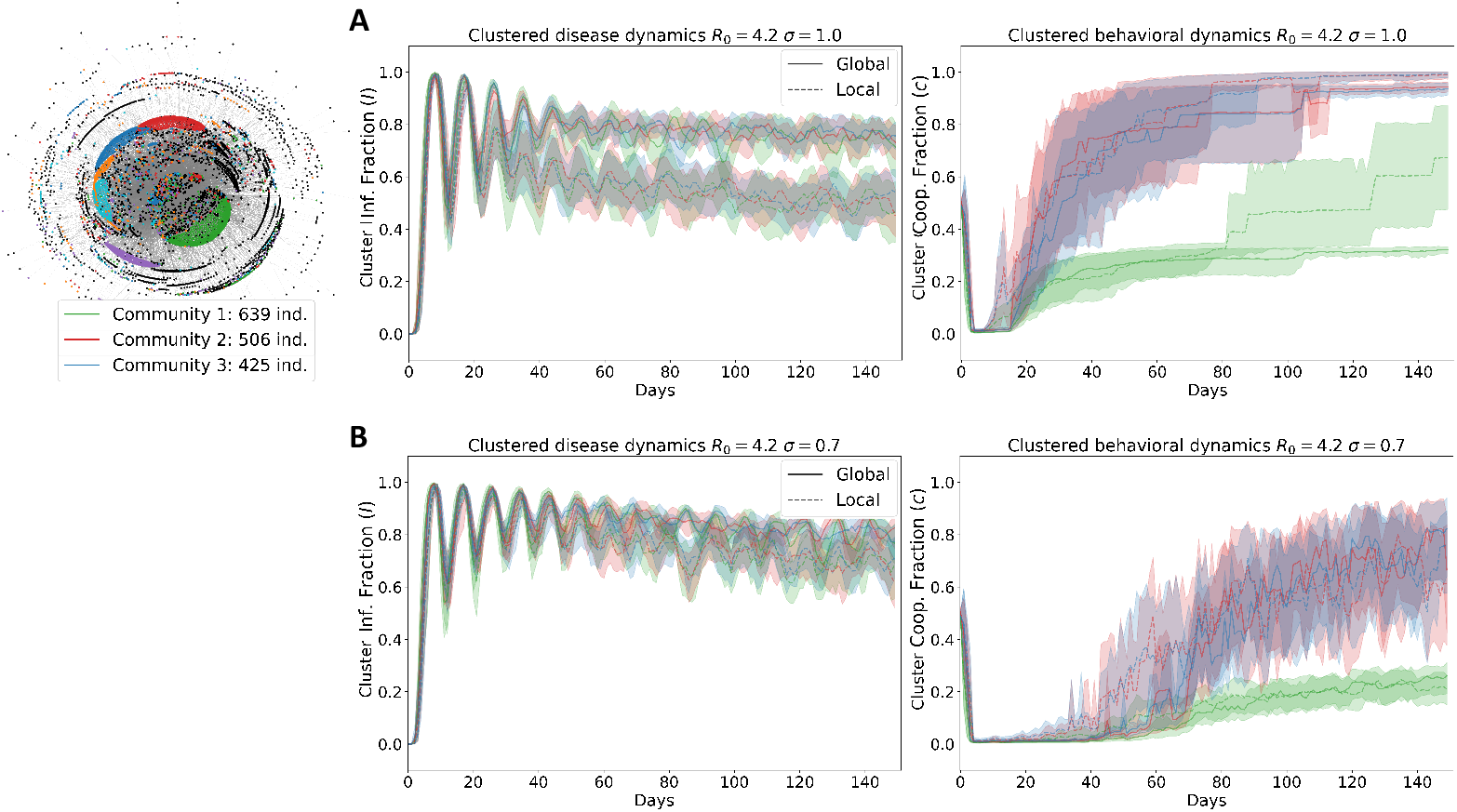
Time dynamics for different levels of consciousness, medium and high consciousness *σ* as described in “Methods” section. Each color represent a hub (set of nodes) identified with the Community clustering algorithm as described in SI. We only studied dynamics in the three hubs with more nodes. For all lines represent the median of the 100 simulations and ribbons represent the 90% quantiles. Left column shows the infected fraction in time *I*(*t*)*/N* and the right column shows the cooperators fraction *c*. The solid line corresponds to running the network model in the global setting while dashed line corresponds to the local setting. We simulate the model for *T* = 150 days after the first infection was seeded.

**Fig 5.**
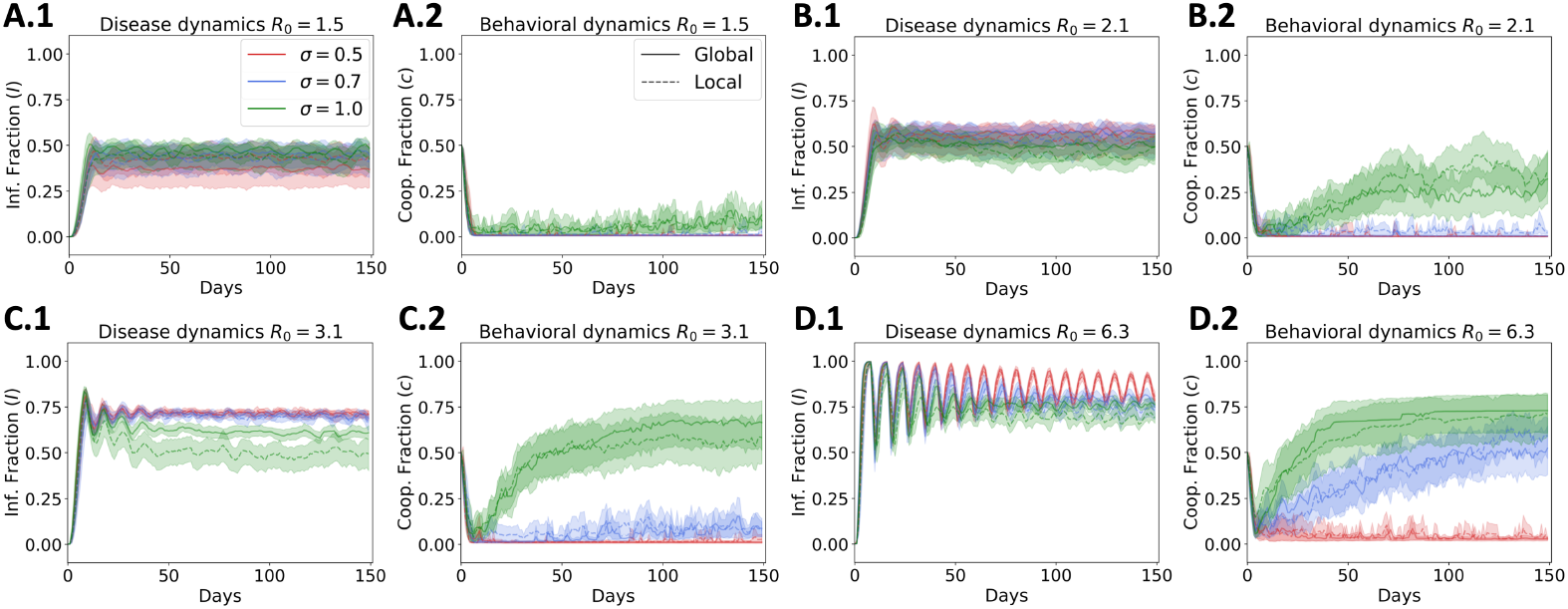
Temporal behavior of scale-free social networks in different contagion scenarios. **A** *R*_0_ = 1.5. **B** *R*_0_ = 2.1. **C** *R*_0_ = 3.1. **D** *R*_0_ = 6.3. Awareness *σ* is varied for 0.5, 0.7 and 1.0 accounting for a scenario of half, partial and full disease awareness, respectively.

**Fig 6.**
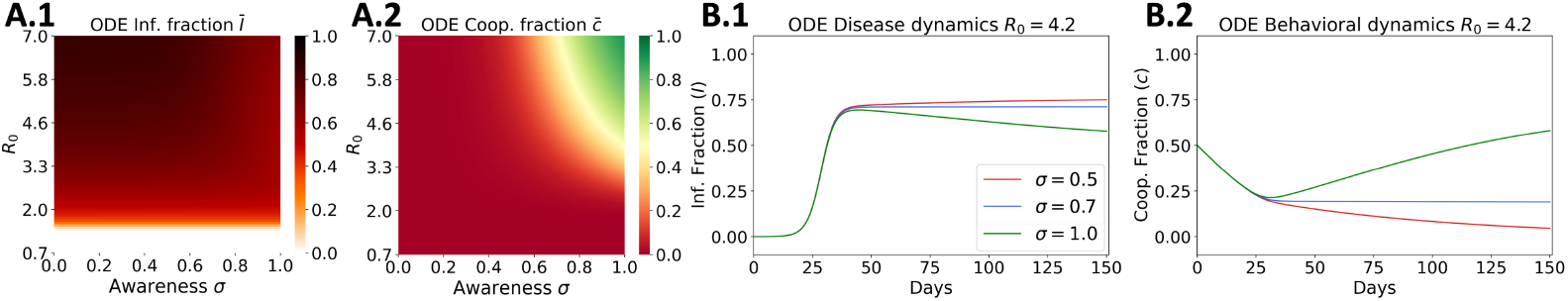
Effect of the basic reproductive number *R*_0_ and awareness *σ* on the infection and cooperation in a mechanistic model. **A** Stable state of infected fraction *Ī* (color coded). **B** Stable state of cooperating fraction 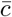 (color coded). **C, D** Temporal dynamics of disease and behavior in a ODE implementation. The basic reproductive number *R*_0_ is fixed to 4.2 and awareness *σ* is varied for 0.5, 0.7 and 1.0 accounting for a scenario of half, partial and full disease awareness, respectively.

Finally, we study the temporal dynamics on a small-world and grid network shown in Fig 7. The temporal dynamics in the ODEs model is shown in Fig. We do not attempt to study clustering structure in the small-world or grid networks as their degree distribution is uniform and therefore the hub might correspond to the whole network. We restrict the analysis of hub dynamics to the scale-free graph.

**Fig 7.**
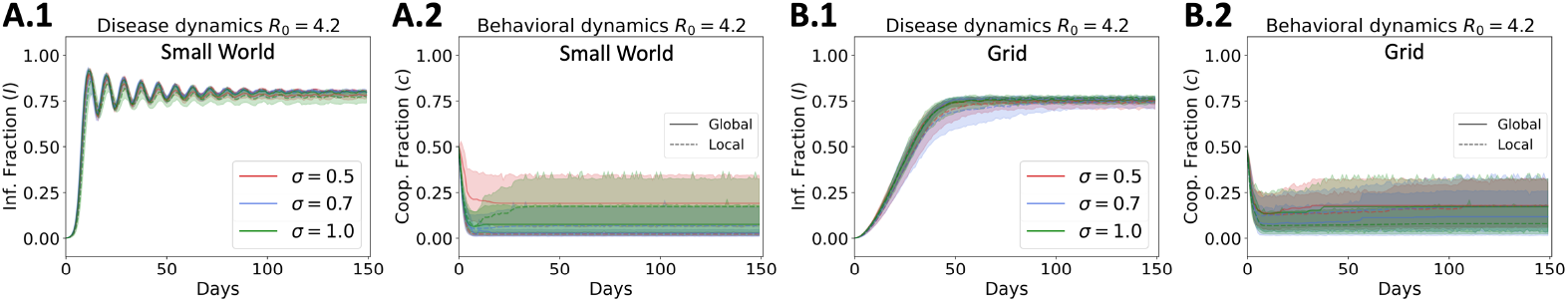
**(Supplementary material)** Temporal behavior of homogeneous social network. **A** Small World graph dynamics for *R*_0_ = 4.2. **B** Grid graph dynamics for *R*_0_ = 4.2. *σ* is varied for 0.5, 0.7 and 1.0 accounting for a scenario of half, partial and full disease awareness, respectively.

## Discussion

Social dynamics play an essential role in the evolution of epidemics and the unfolding of a disease across a population [6, 8]. Specifically, awareness of the state of the epidemic or disease can result in different shapes or patterns in the epidemic curve [9, 27]. The latter implies that the structure of population also plays an important role in both the evolution of disease spread and social dynamics [20]. Real heterogeneous networks such as the scale-free networks are used to resemble real world ones and have been shown to favor cooperation in a social dilemma [20]. We developed a theoretical framework that couples two models typically used for modeling spread of a pathogen in a population the SIS model and for modeling the evolution cooperation in social dilemmas [2, 24]. To our knowledge it is the first infectious disease model to couple social behavior with disease dynamics, using replicator dynamics to examine how awareness of the disease state within a social network may regulate the adoption of protective behaviors. Particularly, we assume social dynamics follow a probability distribution and link the cooperator philosophy to the action toward reducing transmission in the population by assuming contact rate or infection probabilities *β* is a decreasing function of the cooperators individuals in the population.

We have studied the steady state dynamics of the system as shown in Fig 2 and found that ODEs and network model in a scale-free have similar if no identical steady state dynamics as shown in Fig 2A, 2B. As awareness level *σ* in the population increase the final fraction of cooperators almost increase to 100%. Interestingly, these dynamics also exhibit strong relation with disease transmissibility measured here with *R*_0_. We found that for low levels of transmission *R*_0_ *<* 2.1 no matter what level of consciousness is in the population no one cooperates Fig. 2A right and Fig 2B.2 and therefore the final fraction of infected individuals is what the epidemiological system steady state dictates 2A left and Fig 2B.1. These results also confirm our hypothesis that by imposing a discount factor on the human behavior dynamics (here fraction of infected individuals) on the payoff of non-cooperators. The steady state of this dynamics results in a high fraction of the population cooperating towards reducing the disease transmission. This in turn results in a lower fraction of infected individuals when the system reached a steady state.

We found that the steady state of the network model varies greatly depending on the network structure. For the small-world and grid network 2C and 2D we found that the steady state of infected individuals 2C.1 and 2D.1 does not change as a function of the awareness level *σ* of the population. We believe that this behavior emerges as a result of the symmetrical spread of the disease on the graph, also as the degree distribution is uniform across nodes the pathogen spread easily to all the network no matter the initial nodes infected.

Comparing the final fraction of cooperators in the small-world versus the grid graph Fig 2C.2 and Fig 2D.2 respectively, we observed that despite the transmission strength *R*_0_ or the consciousness *σ* nearly 25% of the population cooperates (yellow color in the heat-map). This is confirmed visually by looking at the steady state in Fig. 3 and Fig 7. We hypothesize that these dynamics emerge by the natural structure of the contact network, which naturally pushes the system towards cooperation and sustaining it in time, as has been shown for other real world networks [20].

Our framework accounts dynamically changes in the behavior of an individual, diminishing their own risk of infection as well as the risk of those with whom it interacts [28]. To incorporate this in the models, we considered individual-level contact patterns to capture behavior towards a disease outbreak. This allows an analysis of the disease and behavioral dynamics in complex population structures, and permits explicit demographic predictions that can be studied for public health interventions [29]. We explored the impact of population structure in epidemiological outcomes with different disease and behavioral assumptions set for the R0 and risk awareness.

In general, we have evaluated the effect that communities have over the spread of cooperative strategies towards the mitigation of a disease. We highlight the importance of dense population structures in maintaining cooperative regime, which in turn decreases the epidemic size within the whole population. This crucial characteristic of heterogeneous scale-free graphs is absent in homogeneous scatter and small-world graphs, where this population architecture fail to achieve cooperation in order to counter the spread of an infectious disease.

## Data Availability

This paper do not used any data

## Supporting information

**S1 Fig. Dynamics on Scale Free Networks**.

**S2 Fig. Dynamics on mechanistic - ODEs model**.

**S3 Fig. Dynamics on Small World and Grid network**.

**S4 Algorithm. Clustering Community Algorithm**. This algorithm consists of a two-phase iterative process: **1)** First nodes are separated into different communities, and the gain in modularity *Q*(*H*_*i*_) is calculated each time the nodes are assigned to other *H* communities until there is not a possible positive gain. The process is iterated for all nodes until there is no further improvement; and **2)** the resulting communities are now considered as nodes and a new network is created, described as a refinement stage where the original links of multi-nodes are released and assigned to the same community label [30–32]. Modularity *Q*(*H*_*i*_) for community *H*_*i*_ is defined in Eq. (13) where *d*(*V*) is the total degree of the graph [32, 33].

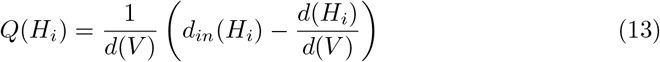

## Acknowledgments

We would like to acknowledge Pallavi Kache, Alejandro Feged-Rivadeneria, Tomas Rodríguez Barraquer and Pablo Cárdenas for their thoughtful comments on the manuscript.

## Notes

### Competing Interest Statement

The authors have declared no competing interest.

### Funding Statement

no external funding was received

### Author Declarations

This paper did not use any data that requires an IRB.

